# Machine Learning-based Clinical Decision Support for Infection Risk Prediction

**DOI:** 10.1101/2023.04.27.23289212

**Authors:** Ting Feng, David Noren, Chaitanya Kulkarni, Sara Mariani, Claire Zhao, Erina Ghosh, Dennis Swearingen, Joseph Frassica, Daniel McFarlane, Bryan Conroy

**Affiliations:** Philips Research North America, Cambridge MA, USA; Philips Research Bangalore, Bengaluru, India; Department of Medical Informatics, Banner Health, Phoenix AZ, USA; Department of Biomedical Informatics, University of Arizona College of Medicine, Phoenix AZ, USA; Institute for Medical Engineering and Science, Massachusetts Institute of Technology, Cambridge MA, USA

**Keywords:** Healthcare-associated infection (HAI), Machine Learning, Clinical Decision Support (CDS)

## Abstract

**Background:** Healthcare-associated infection (HAI) remains a significant risk for hospitalized patients and a challenging burden for the healthcare system. This study presents a clinical decision support tool that can be used in clinical workflows to proactively engage secondary assessments of pre-symptomatic and at-risk infection patients, thereby enabling earlier diagnosis and treatment.

**Methods:** This study applies machine learning, specifically ensemble-based boosted decision trees, on large retrospective hospital datasets to develop an infection risk score that predicts infection before obvious symptoms present. We extracted a stratified machine learning dataset of 36,782 healthcare-associated infection patients. The model leveraged vital signs, laboratory measurements and demographics to predict HAI before clinical suspicion, which is defined as the order of a microbiology test or administration of antibiotics.

**Results:** We find that our best performing infection risk model achieves a cross-validated AUC of 0.88 at 1-hour before clinical suspicion and maintains an AUC>0.85 for 48-hours before suspicion by aggregating information across demographics and a set of 163 vital signs and laboratory measurements. A second model trained on a reduced feature space comprising demographics and the 36 most frequently measured vital signs and laboratory measurements can still achieve an AUC of 0.86 at 1-hour before clinical suspicion. These results compare favorably against using temperature alone and clinical rules such as the quick Sequential Organ Failure Assessment (qSOFA) score. Along with the performance results, we also provide an analysis on model interpretability via feature importance rankings.

**Conclusions:** The predictive model aggregates information from multiple physiological parameters such as vital signs and laboratory measurements to provide a continuous risk score of infection that can be deployed in hospitals to provide advance warning of patient deterioration.

## BACKGROUND

Healthcare-associated infection (HAI), also referred to as nosocomial infection, remains a significant risk for hospitalized patients and a significant burden on healthcare systems. It has been reported that approximately 1 in 31 hospital patients develop an HAI on any given day [1], and nearly 99,000 people in the U.S. die annually from HAIs [2]. Recent data shows that the incidence of HAI’s increased during the pandemic (2020) revealing the fragile nature of interventions aimed at prevention [3]. Over the last decade, the CDC has developed guidelines and strategies for the prevention of HAIs, focusing on improving clinical practice and antibiotic stewardship. While this guidance has shown some utility in lowering the incidence across several types of HAI, improving the outcomes for those who become infected remains challenging, particularly for the critically ill.

Early detection of de-novo infectious disease is critical for improving the outcomes of infected patients [4] [5], for the timely implementation of control measures critical to preventing its spread [6], and for reducing substantial healthcare cost associated with preventable HAIs [7]. Hospitalized patients suffering from influenza, up to 20% of whom are nosocomial in origin, have better outcomes when treated with antiviral agents immediately after symptoms present [8]. Antibiotic treatment has also been shown to be more effective in producing better outcomes for sepsis patients when administered early in the progression of the infection, particularly for mechanically ventilated patients [4] [5].

Clinical decision support (CDS) tools have received a great deal of attention over the last decade, including those focused on the detection of infection [9] [10] [11]. Many of these CDS tools are rule based and developed through physician consensus and guidelines. These include more standardized solutions like the Acute Kidney Injury (AKI) eAlert that has been deployed in hospitals in Wales [12] [13] and the National Early Warning Score (NEWS) score that is standard for detecting general clinical deterioration in the UK [14]. While these approaches benefit from clinician experience, they are simplified to remain generalizable and fail to capture the complete clinical context required to discriminate difficult or atypical cases. In addition, these approaches are not easily tailored or adapted, for example, to specific patient populations. More recently, several studies have suggested data-driven approaches to create physiological risk prediction algorithms, including in the areas of infection and sepsis prediction [9] [15] [16] [17].

This study uses machine learning applied on large retrospective hospital datasets to develop a clinical decision support (CDS) algorithm for the early detection of infection in hospitalized patients. By aggregating information across demographics and a set of 163 vital signs and laboratory measurements, we find our best-performing model can achieve a cross-validated AUC of 0.88 at 1-hour before clinical suspicion, and maintains an AUC>0.85 for the 48-hour period prior to clinical suspicion of infection. By distilling the model down to a set of 36 most frequently measured vital signs, laboratory measurements and demographics, we can still maintain an AUC of 0.86 at 1-hour before clinical suspicion. In the results, we further contrast our models against established clinical scoring systems – quick Sequential Organ Failure Assessment (qSOFA), and against tracking individual vital signs alone (e.g., temperature, etc.).

## METHODS

### Description of data

We combined clinical data from three large hospital datasets: the MIMIC-III (Medical Information Mart for Intensive Care III) database collected from 2001 to 2012 [18], the eICU dataset from Philips’ electronic ICU telemedicine business collected from 2003 to 2016 [19], and a dataset of electronic medical records from Banner Health collected from 2010 to 2015. In total, the combined dataset includes over 6.5 million patient encounters collected from more than 450 hospitals. Supplemental Figure 1 indicates the types of data present in each hospital dataset.

### Ethical Approval

The MIMIC-III project was approved by the Institutional Review Boards of Beth Israel Deaconess Medical Center (Boston, MA) and the Massachusetts Institute of Technology (Cambridge, MA). Use of the eICU data was approved by the Philips Internal Committee for Biomedical Experiments. Banner Health data use was a part of an ongoing retrospective deterioration detection study approved by the Institutional Review Board of Banner Health and by the Philips Internal Committee for Biomedical Experiments. Requirement for individual patient consent was waived because the project did not impact clinical care, was no greater than minimal risk, and all protected health information was removed from the limited dataset used in this study.

### Infection and control cohort extraction

We define infection patients as those who 1) have a confirmed infection diagnosis, and 2) have data indicating clinical suspicion of infection. Patients in the infection cohort were selected as those with confirmed infection diagnoses via ICD-9 and whose timing of clinical suspicion of infection could be localized by a microbiology culture test order. Infection patients were then further screened into an HAI cohort if the timing of clinical suspicion of infection occurred at least 48 hours after admission. Patients in the control cohort were selected as those who have neither an infection-related ICD-9 diagnosis code nor any microbiology culture tests ordered. Since the selection criteria identified a much larger set of control patients than HAI patients, we down-sampled the control cohort population to maintain a prior infection odds (prevalence) of 12.5%. This ensured that the training dataset would not be overly dominated by control patients, while still maintaining the HAI cohort as the minority class. Figure 1 shows the general decision scheme behind this methodology. Curation of infection ICD-9 codes is described in detail in the Supplementary Materials.

**Figure 1:**
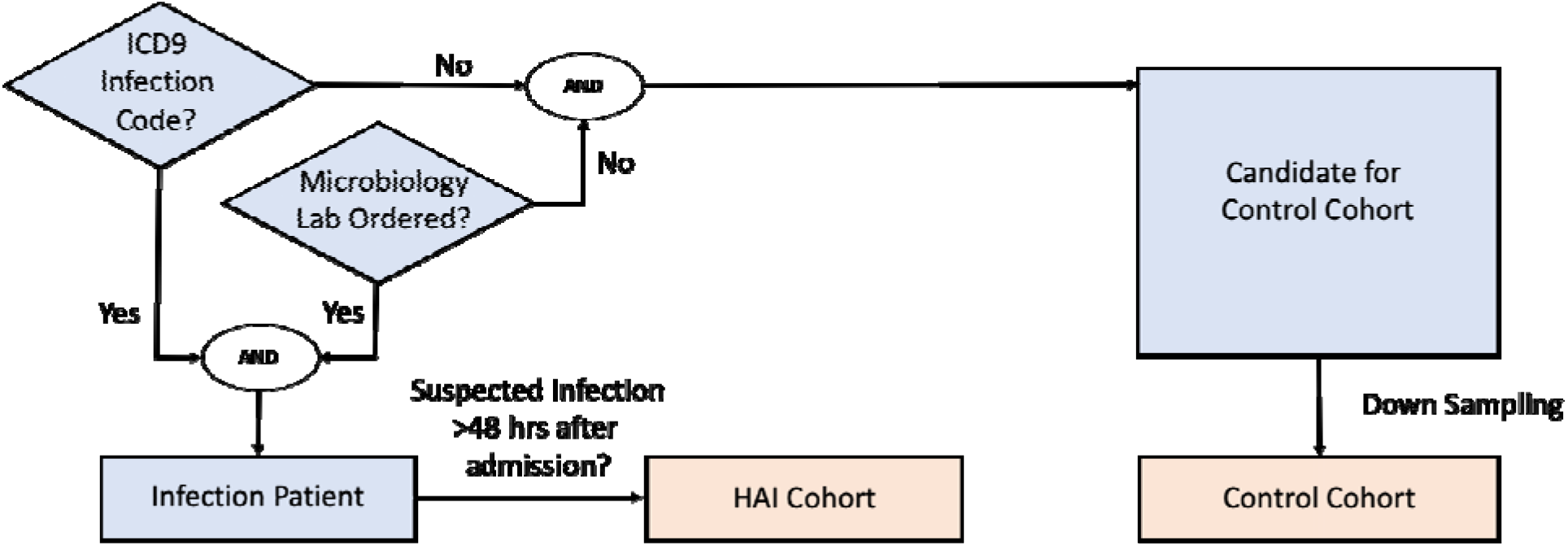
Cohort inclusion/exclusion criteria flow diagram

For a minority of hospitals, microbiology charting data was either missing, sporadic, or incomplete. In such cases, the microbiology culture test criterion was replaced with administration of non-prophylactic antibiotics. The cohort selection was otherwise the same: infection patients were those with at least one administration of non-prophylactic antibiotics and who had at least one ICD-9 code indicating infection, while control patients were selected as those who had neither an ICD-9 code nor any administration of non-prophylactic antibiotics.

Clinical suspicion of infection (and screening for the HAI cohort) was then derived using the administration time of first non-prophylactic antibiotics. We validated, in the MIMIC-III dataset, that the two criteria (microbiology culture test versus non-prophylactic antibiotics administration) yield a large overlap of the selected cohorts (see Supplementary Materials). Extraction of antibiotic records and non-prophylactic labelling details are also described in the Supplementary Materials.

For control patients, we generated a synthetic event time, such that clinical data used for prediction could be extracted in the same way as was done for the infection patients. To reduce bias, and to ensure sufficient data prior to event time for model building, we randomly assigned a time-point that is at least 48 hours after the patient’s first clinical measurement, and that precedes the end of the patient’s hospital stay as the synthetic event time.

### Description of features and feature subsets used by the models

The extracted features are comprised of three sets of information: demographics (e.g., age, gender, height, weight), vital sign measurements (e.g., heart rate, blood pressure, temperature), and laboratory measurements (e.g., metabolic panels, complete blood count, and arterial blood gas). After feature extraction from each of the three hospital datasets, we applied an extensive preprocessing and cleaning pipeline to create a common and consistent dataset. A full list of the features is given in the Supplemental Materials in Table A1.

For the purpose of training our machine learning algorithms, we defined an observation time as one hour before each patient’s clinical suspicion of infection (or randomly assigned event time for control patients). We then extracted the latest measured value of each feature leading up to the observation time and assembled these measurements into a physiological state vector for each patient. This feature vector was then augmented with features characterizing temporal trends from vital sign measurements during the 48-hour window preceding the observation time. To mitigate sensitivity to outliers, we applied physiologic plausibility filters to the vital signs before calculating trends. Trend features on laboratory measurements were excluded since they tend to be measured aperiodically (e.g., daily). We extracted five trend features for the for vital signs: Temperature, Heart Rate, Systolic, Diastolic, and Mean Blood Pressures, Oxygen Saturation^1^ (SpO2), and Respiration. For example, these trend features for Heart Rate are: Avg(Heart Rate): The average heart rate value over a 48-hour window

- Min(Heart Rate): The minimum heart rate value over a 48-hour window
- Max(Heart Rate): The maximum heart rate value over a 48-hour window.
- Var(Heart Rate): The variance of heart rate over a 48-hour window
- CoefVar(Heart Rate), or CV(Heart Rate): The coefficient of variation of heart rate over a 48-hour window, defined as the standard deviation divided by the mean

During the validation stage of our algorithm, we additionally applied the classifiers trained on the one-hour before observation time to earlier time windows in order to characterize predictive performance over time. In those instances, we extracted a physiological state vector at earlier observation times in an analogous manner. Figure S3 provides a visual summary of the feature extraction pipeline.

### Description of algorithms used

We employed two groups of algorithms: (a) linear classifiers, which identify a separating hyperplane in the original feature space; and (b) ensemble-based methods, which iteratively construct a powerful classifier from a set of “weak” nonlinear classifiers. We chose linear classifiers and ensemble-based methods over neural network techniques because we preferred to maintain interpretability of the trained model for clinical deployment, and to minimize the usage of computation resources to enable flexible applications. For linear classifiers we choose logistic regression, and for ensemble methods we benchmarked against Abstained Adaptive Boosting with univariate decision stumps [20] and Gradient Boosting of decision trees using the XGBoost algorithm [21]. Since our dataset is imbalanced in terms of infection prevalence, we employed stratified cross-validation, and we did this for each of the three hospital datasets separately: with stratification, both the ratio of control to infection patients, and the ratio of patients from different hospital datasets are maintained in both training and testing sets. Information about imputation, hyperparameter tuning and performance evaluation is detailed in the Supplemental Materials.

### Description of model interpretation methods

The Adaptive Boosting algorithm with decision stumps can be expressed as a generalized additive model of the form where R(x) is the composite (ensemble) classifier, x_1_,x_2_,…,x_p_ are the p feature inputs, and r_j_(x_j_), j=1,…,p are the “weak learner” classifiers learned for each feature. In this case, infection patients are labeled as class 1 (controls are class -1), so that a larger value of R(x) indicates the classifier’s stronger confidence of the patient having infection. As a result, each r_j_(x_j_) can be interpreted as an infection risk function evaluated with respect to a single feature. In order to control for the impact of feature missingness, we analyzed the relative importance of features through each r_j_(x_j_) in two ways: (1) *total feature importance*, which evaluates a feature’s importance across the entire cohort; and (2) *adjusted feature importance*, which isolates the feature’s contribution on the subset of patients that have the feature measured. Therefore, *total feature importance* gives an indication of a feature’s effectiveness under typical hospital workflow conditions, while *adjusted feature importance* can identify discriminative features despite being less frequently measured.

The Gradient Boosting algorithm can be interpreted using SHAP (Shapley Additive exPlanations) method [22]. SHAP assigns each feature an importance value for a particular prediction, therefore we can compare feature importance by examining the distribution of SHAP values which represent the impacts each feature has on the model output.

## RESULTS

The cohort selection criteria resulted in a total training dataset size of 293,109 patients (256,327 control patients; 36,782 HAI patients). Of these patients, 63% are from the Banner Health dataset, 32% are from the eICU dataset, and 5% are from the MIMIC-III dataset. The majority of these patients are treated under ICU or general ward settings. Between the two infection cohort criteria (microbiology culture orders vs non-prophylactic antibiotics administration), 26,599 HAI patients are identified from microbiology lab and ICD-9 code, while 10,183 infection patients are identified from non-prophylactic antibiotic administration and ICD-9 code.

### Model performance

We compared machine learning algorithms in their ability to discriminate infection from control patients using clinical data acquired up to one hour before clinical suspicion of infection. Our results show that gradient boosting with two level decision trees yielded the best performance with a mean AUC of 0.88, Specificity of 0.93 and Sensitivity of 0.54 at the break-even point (where Sensitivity is approximately equal to positive predictive value (PPV), see Supplemental Materials), Sensitivity of 0.80 and 0.64 respectively for when Specificity is 0.80 and 0.90 (Table 1: Xgboost). Abstained Adaptive Boosting with decision stump achieved a mean AUC of 0.85, Specificity of 0.92 and Sensitivity of 0.47 at break-even point, Sensitivity of 0.73 and 0.54 respectively for when Specificity is 0.80 and 0.90 (Table 1: Abstained AdaBoost). Logistic regression performs poorly compared with ensemble algorithms, with a mean AUC of 0.77, Specificity of 0.91 and Sensitivity of 0.40 at break-even point, Sensitivity of 0.60 and 0.43 respectively for when Specificity is 0.80 and 0.90 (Table 1: Logistic Regression). These results suggest that ensemble models are superior to linear models in predicting infection.

**Table 1:** Performance of infection prediction at one hour before clinical suspicion of infection.

| **Algorithm** | **AUC** | **Sensitivity (Spec)**<br>**Break-Even Point** | **Sensitivity**<br>**@ Specificity=0.8** | **Sensitivity**<br>**@**<br>**Specificity=0.9** |
| --- | --- | --- | --- | --- |
| GradientBoost | 0.884 | 0.537 (0.934) | 0.800 | 0.635 |
| Abstained<br>AdaBoost | 0.852 | 0.469 (0.924) | 0.731 | 0.536 |
| Logistic<br>Regression | 0.772 | 0.399 (0.914) | 0.597 | 0.431 |
| GradientBoost –<br>exclude lab | 0.810 | 0.415 (0.916) | 0.622 | 0.449 |
| GradientBoost –<br>reduced features | 0.862 | 0.499 (0.928) | 0.750 | 0.574 |

Next, we asked if ensemble models perform better than established empirical rules and clinical scores in infection prediction. First, fever or high body temperature (>98.6 F) is one of the first symptoms that lead to clinical suspicion of infection. Therefore, we compared temperature measurements between the infection and control cohorts, and calculated the discriminative power of temperature at one hour before infection suspicion. Temperature by itself has an AUC = 0.59 for detecting infection, which is far inferior to performance achieved with gradient boosting (AUC = 0.88). Second, qSOFA – quick Sequential Organ Failure Assessment – was introduced by the Third International Consensus Definitions for Sepsis and Septic Shock task force in 2016, and is proposed as a quick assessment tool for identifying sepsis among patients with infection [23]. Based on the Sepsis-3 criteria, we extracted Glasgow Coma Score, Systolic Blood Pressure, and Respiratory Rate from the medical database, and derived qSOFA scores at one hour before clinical suspicion of infection. In total 111,651 qSOFA scores were extracted, 22,460 from infection cohort and 89,191 from control cohort (infection prevalence = 20.1%). We then calculated the area under ROC curve of infection prediction by using qSOFA alone. qSOFA by itself has an AUC = 0.59 when predicting infection at one hour before suspicion of infection. To ensure a fair comparison with ensemble models, we re-trained the Gradient Boosting algorithm using data from the subset of patient cohort that have qSOFA available. Gradient Boosting on the patient subset achieves an AUC of 0.83 which is substantially better than the performance of qSOFA. Overall our results suggest advantages of ensemble models over established clinical methods in infection prediction.

We further benchmarked ensemble model performance when feature sets are reduced. First, we excluded all lab measurements and focused on 14 vital signs and demographics factors (plus 50 derived trend features), as they are continuously available and more predictably available than lab measurements. Gradient Boosting, re-trained from the feature space excluding labs, achieved a mean AUC of 0.81, Specificity of 0.92 and Sensitivity of 0.42 at break-even point, Sensitivity of 0.62 and 0.45 respectively for when Specificity is 0.80 and 0.90 at one hour before clinical suspicion of infection (Table 1: GradientBoost – exclude lab). Second, we excluded infrequently measured features that are available for less than 70% of the patient cohort. This produced a reduced feature space with 36 vitals, demographics and laboratory measurements (plus 32 derived trend features). Gradient Boosting model, re-trained from frequently measured features, achieved a mean AUC of 0.86, Specificity of 0.93 and Sensitivity of 0.50 at break-even point, Sensitivity of 0.74 and 0.57 respectively for when Specificity is 0.80 and 0.90 at one hour before clinical suspicion of infection (Table 1: Xgboost – reduced features). These results suggest that it is possible to obtain good performance when reducing the total feature space by half.

In addition, we investigated the infection prediction performance of ensemble models at earlier time points. We applied the most interpretable model (Abstained AdaBoost) and the best performing model (Gradient Boosting) to earlier observation windows to characterize predictive performance over time using the full feature space (Figure 2). Despite degraded model performance over time, Gradient Boosting maintains an AUC>0.85, while Adaptive Boosting maintains an AUC>0.81 for 48 hours before clinical suspicion. These results support an assertion that it is possible to predict hospital acquired infection earlier, up to 48 hours before clinical suspicion of infection.

**Figure 2:**
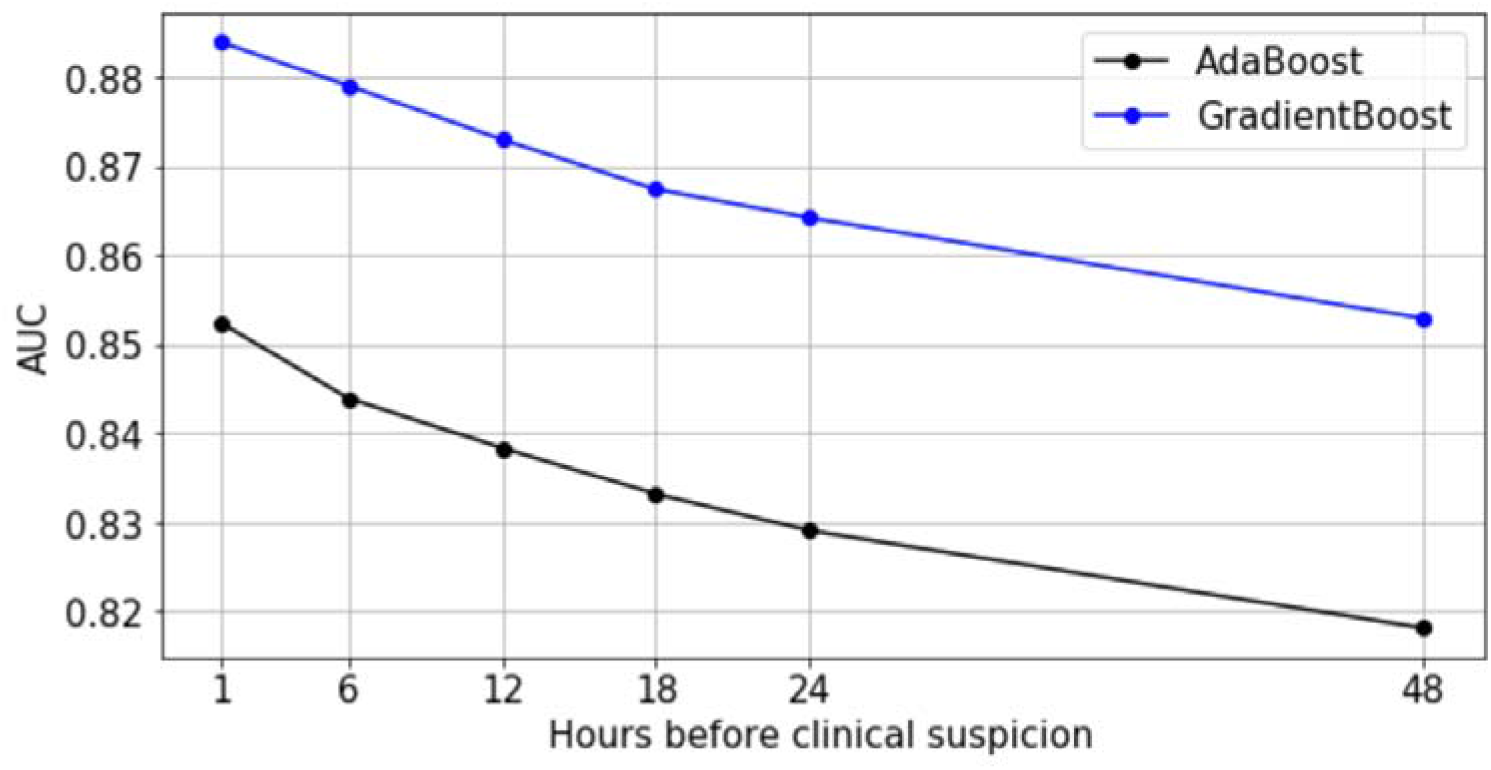
Predictive performance of AdaBoost and GradientBoost models relative to time of clinical suspicion

### Model interpretation

To better understand the biomarkers leveraged by the ensemble-based models, we first analyze the AdaBoost algorithm with decision stumps since it is easier to interpret, and then contrast with feature importance scores on the GradientBoost algorithm with decision trees using the SHAP (Shapley Additive exPlanations) method [22].

We first examined the top 15 features ranked by *total feature importance* and *adjusted feature importance* derived from Abstained Adaptive Boosting model trained in the full feature space. (Table 2). As described in Methods, *total feature importance* evaluates a feature’s importance across the entire cohort, and *adjusted feature importance* isolates the feature’s contribution on the subset of patients that have the feature measured. From both metrics, we found that the top 15 features are a mix of laboratory measurements and vital signs. Adjusted feature importance, in particular, identifies discriminative features from laboratory measurements despite being less frequently measured.

**Table 2:** Feature Importance Rankings from Abstained AdaBoost model (top 15). Total feature importance evaluates a feature’s importance across the entire cohort; adjusted feature importance isolates the feature’s contribution on the subset of patients that have the feature measured.

| Total Feature Importance |  | Adjusted Feature Importance |  |
| --- | --- | --- | --- |
| <u>Rank</u> | <u>Feature</u> | <u>Rank</u> | <u>Feature</u> |
| 1 | Albumin | 1 | Albumin |
| 2 | Max(SpO2) | 2 | TIBC |
| 3 | pH | 3 | Fibrinogen |
| 4 | Min(SpO2) | 4 | Temperature |
| 5 | Temperature | 5 | ESR |
| 6 | Avg(SpO2) | 6 | PVRI |
| 7 | Var(SpO2) | 7 | Max(Temperature) |
| 8 | Lactate | 8 | Urinary RBC |
| 9 | Bands | 9 | Avg(Respiration) |
| 10 | Max(Temperature) | 10 | WBC |
| 11 | Avg(Respiration) | 11 | BUN |
| 12 | CV(SpO2) | 12 | CRP |
| 13 | FiO2 | 13 | Ferritin |
| 14 | WBC | 14 | Neutrophils |
| 15 | Bicarbonate | 15 | Var(Temperature) |

The learned risk functions behave in clinically interpretable ways. Figure 3 visualizes the risk functions (black) for a subset of the most important laboratory features, along with population distribution underlays for infection (red) and control (blue) populations. The learned risk functions for these representative features are either monotonically increasing, suggesting that an elevation of the respective clinical measurement is associated with higher infection risk; or monotonically decreasing, suggesting that a decrease of the respective clinical measurement is associated with higher infection risk. During training, each risk function is assembled from a collection of decision stumps that identify key feature thresholds that distinguish levels of infection risk. The scale of the risk function (the y-axis in Figure 3 plots) is unitless, but can be used to compare the relative importance of features (see Table 2 for further details on feature importance).

**Figure 3:**
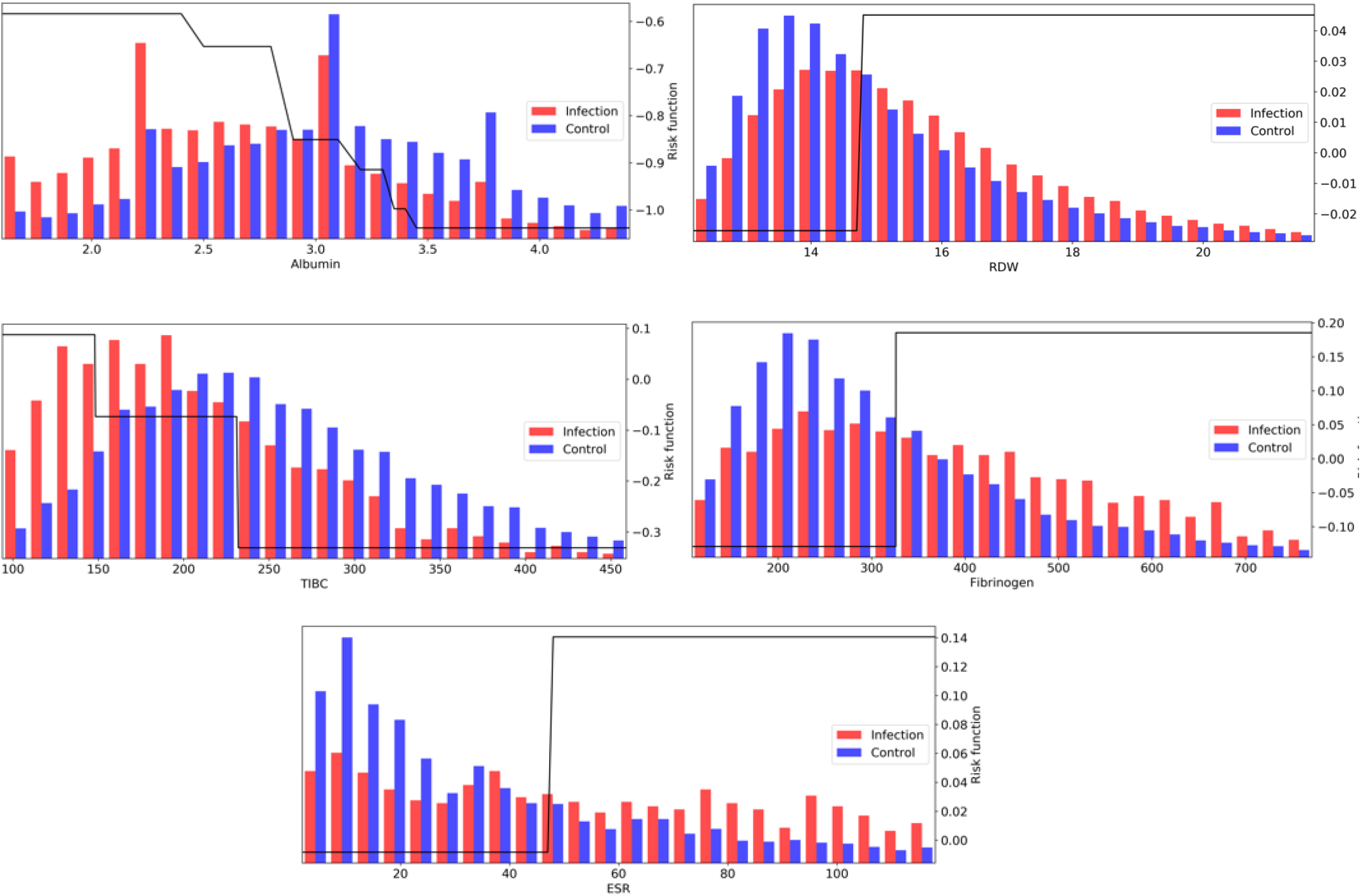
AdaBoost risk functions (black) for a subset of the most important laboratory measurements, along with population distribution underlays for infection (red) and control (blue) populations

Amongst laboratory measurements, a number of features associated with, but not necessarily specific to, inflammation were identified. The top feature across both scoring metrics was associated with hypoalbuminemia (low albumin levels < 3 g/dL), which has been shown to correlate with inflammation, shock, and sepsis [24]. High RDW (>15%) was also a strong biomarker, with literature showing it correlated with inflammation markers CRP and ESR [14]. With respect to the *adjusted feature importance* score, a number of infrequently measured features, but highly discriminative, were identified by the model, all of which show associations with inflammatory response: low TIBC (<240 mcg/dL; prevalence=3%), elevated Fibrinogen (>325 mg/dL; prevalence=5%), and elevated ESR (> 45 mm/hr; prevalence=2%).

Many other laboratory values were also discriminative. Increased risk is identified when Bicarbonate levels fall below approximately 24 mEq/L, which may be indicative of metabolic acidosis, in particular lactic acidosis (elevated Lactate levels above 1.5 mmol/L were also contributing to infection risk). White blood cell concentrations were also strong indicators in the top 15 features, with elevated Bands and Neutrophil concentrations [25]. Other notable indicators are low HDL and LDL cholesterol levels [26], and increases in blood platelets, which is a sign of host defense and induction of inflammation and tissue repair in response to infection onset [27].

Although laboratory measurements play a significant role, the model also aggregates information from a number of vital signs. The infection risk function based on temperature increases rapidly above 37.8C, although this accounts for a small percentage of infection patients (5105 out of 40406 (∼12.6%) of infection patients registered a fever >= 37.8C at the 1-hour window). For controls, 5579 out of the 96505 control patients (∼5.8%) exhibited a fever >= 37.8C. Infection patients tend to have an elevated heart rate and macro variability, which is reported to be critical for the diagnosis and prognosis of infection by many studies [28] [29]. For blood pressure, patients tend to have a decreased blood pressure (systolic, diastolic, and mean), and this effect was often selected by the classifier. Many trend variability features on vitals were selected across temperature, heart rate, blood pressure, oxygen saturation(SpO2), and respiration, as the infection cohort tends to exhibit a heavier right tail in feature variance measures. Changes in vital signs are also reported in the literature to accompany the development of infection [30] [31].

We additionally applied SHAP analysis to extract feature importance rankings from the Gradient Boosting method (Figure 4). We have observed overlaps in the selected features between the more interpretable AdaBoost model and Gradient Boosting, such as Albumin, SpO2, Bicarbonate, Temperature, Lactate and BUN.

**Figure 4:**
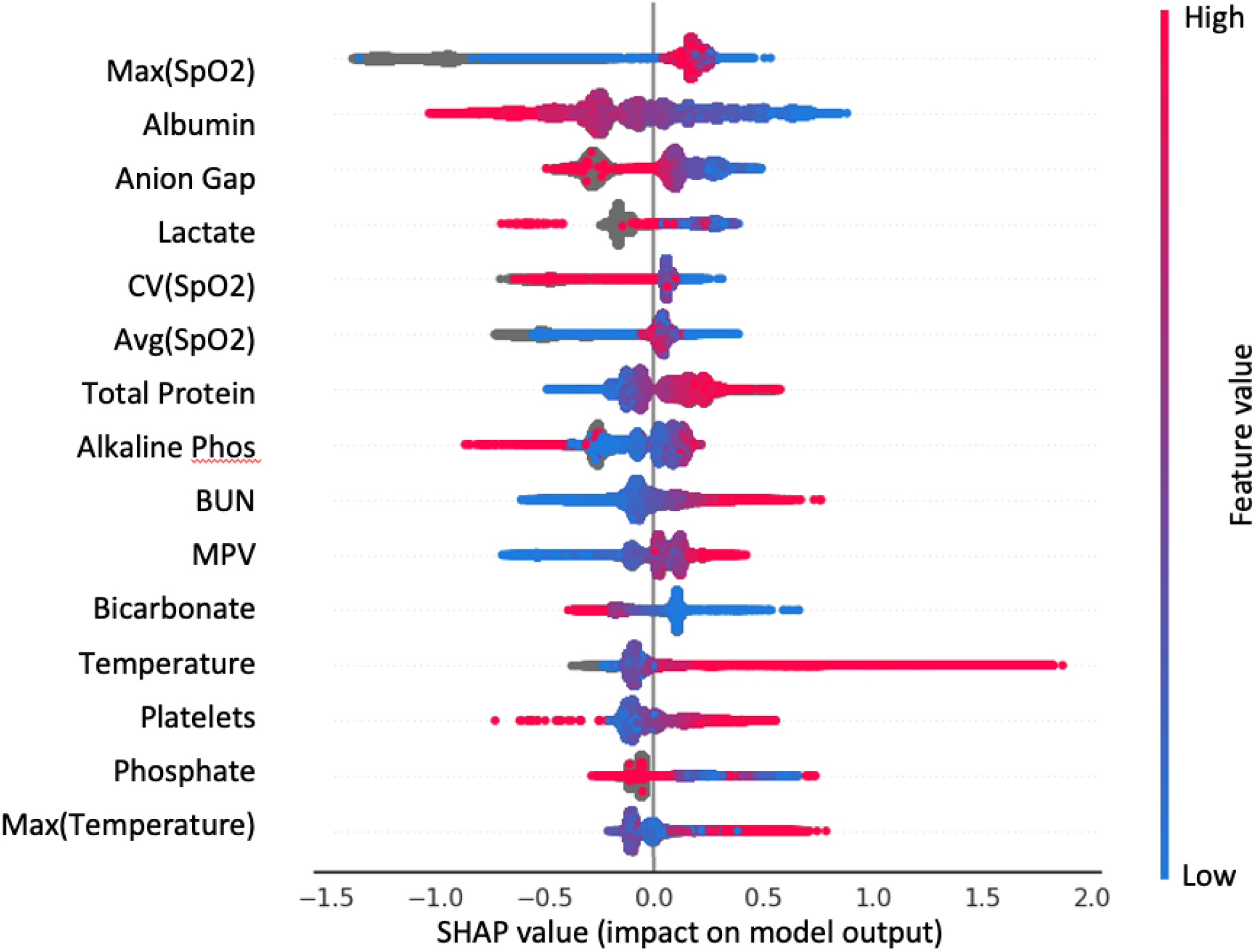
Top 15 important features of the GradientBoost model from SHAP analysis. Each dot is a patient; color indicates the value of the feature. SHAP value is on the x-axis: large positive value - feature contributes strongly to predict infection; large negative value – feature contributes strongly to predict control

### Algorithm performance on infection subgroups

Patients’ host responses to pathogens vary between pathogens and primary sites of infection which result in heterogeneous physiological changes. The extracted HAI cohort is mainly from, ranked by high to low prevalence, the following five infection types (defined by ICD-9 codes - see Supplementary Materials): pneumonia (17,224 patients), bloodstream infection (12,891 patients), bone/joint/tissue/soft tissue infection (11,613 patients), sepsis (9,643 patients) and urinary system infection (9,118 patients). Note that these patients are primarily from ICUs or general wards, and some patients can have more than one HAI. To compare detection performance on different infection types, we calculated recall (Sensitivity) from the model for patient subgroups of different infection types (Figure 5). We found that the infection model (Table 1: Xgboost) has the highest recall in predicting Sepsis (recall = 0.70) and bloodstream infection (recall = 0.67), followed by pneumonia (recall = 0.61), bone/joint/tissue/soft tissue infection (recall = 0.50) and urinary system infection (recall = 0.46). This result indicates that the infection model performs the best in predicting subgroups of patients that have high acuity.

**Figure 5:**
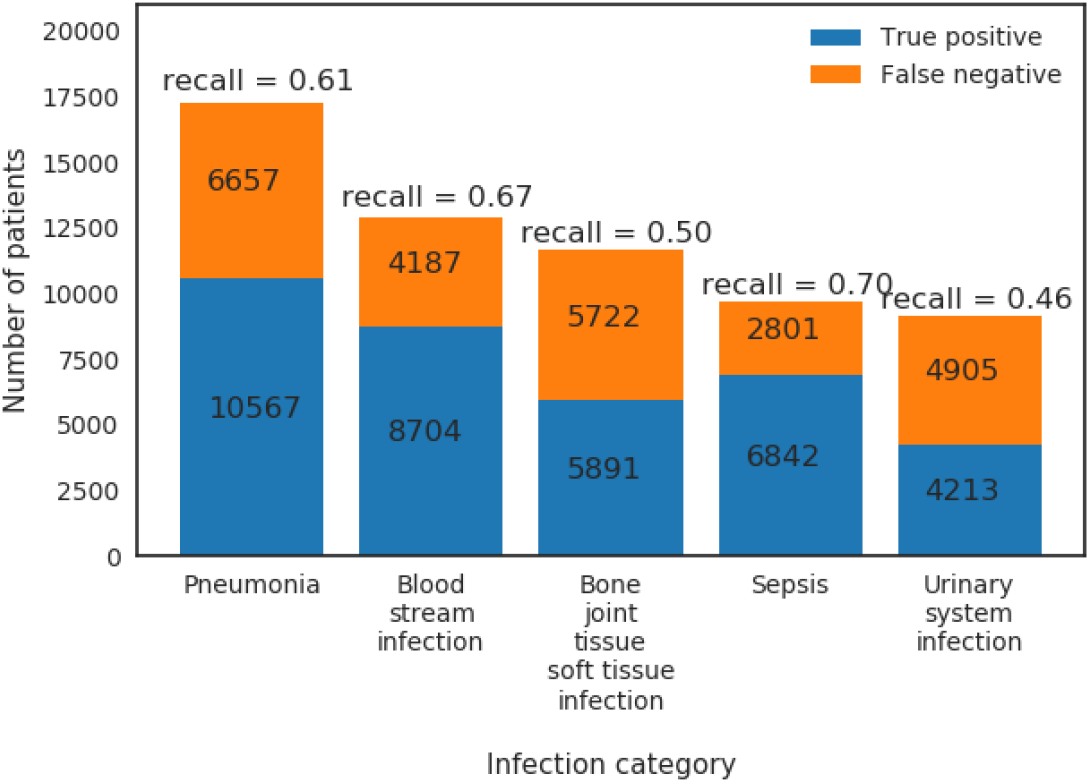
True positives and false negatives from GradientBoost model for the top five prevalence infection categories

### Impact of comorbidities on algorithm performance

The previous section assessed true positive rates (recall/sensitivity) for various infection types. By the same token, we may also characterize true negative performance of the algorithm with respect to various chronic comorbidities exhibited by the control patient population. To do so, we calculated the Elixhauser Comorbidity Index [32] for each control patient, which associates diagnostic ICD-9 codes (see Table 2 of [32]) with a set of 30 comorbidity categories. Of the 256,327 control patients, 194,364 (76%) exhibited at least one comorbidity – see Figure 6 for a summary of prevalence of each comorbidity category amongst control patients. We then calculated the infection model’s true negative rate (TNR) on the control patient population that exhibited each of the 30 comorbidity categories. In addition, we compared true negative rate for control patients with at least one comorbidity (76% of all control patients, labeled “With comorbidities”) to the true negative rate for control patients without any documented comorbidities (24% of all control patients, labeled “Without comorbidities”) – see Figure 7.

**Figure 6:**
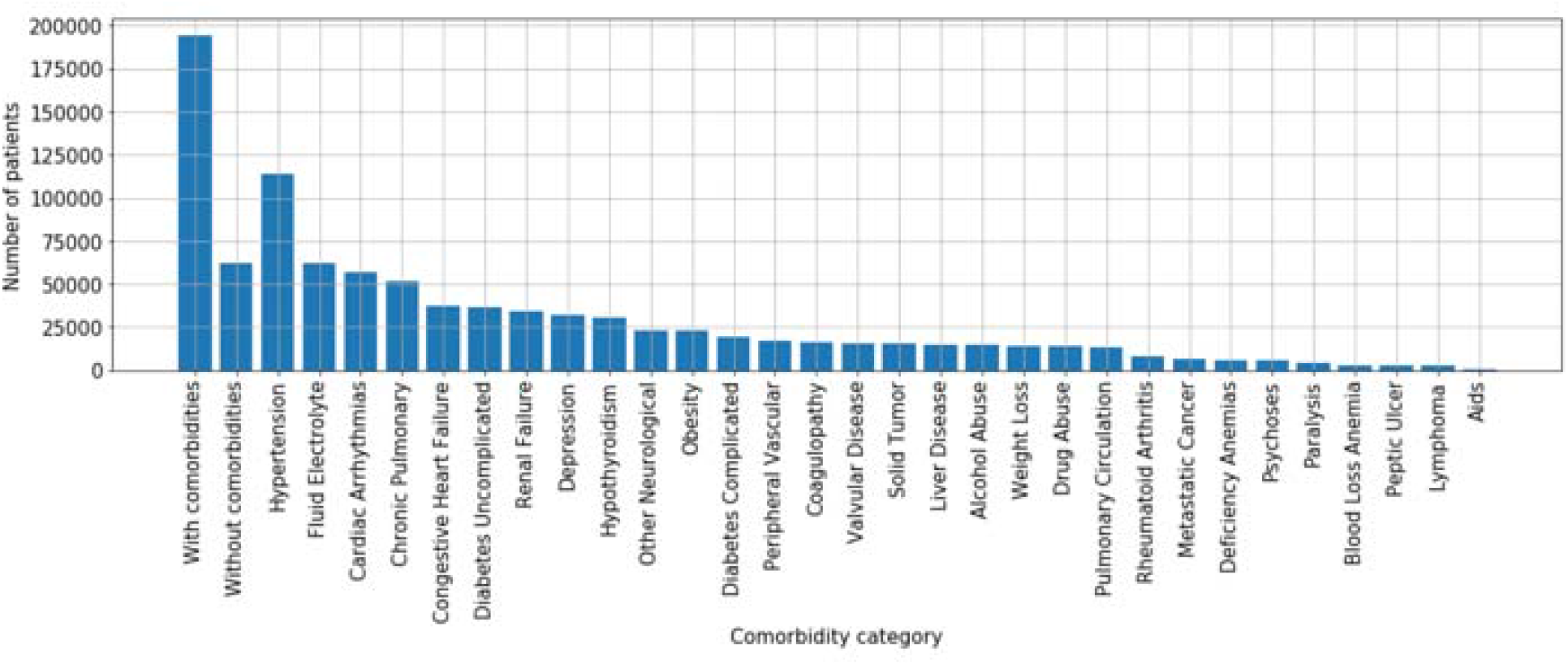
Comorbidity prevalence amongst control patients

**Figure 7:**
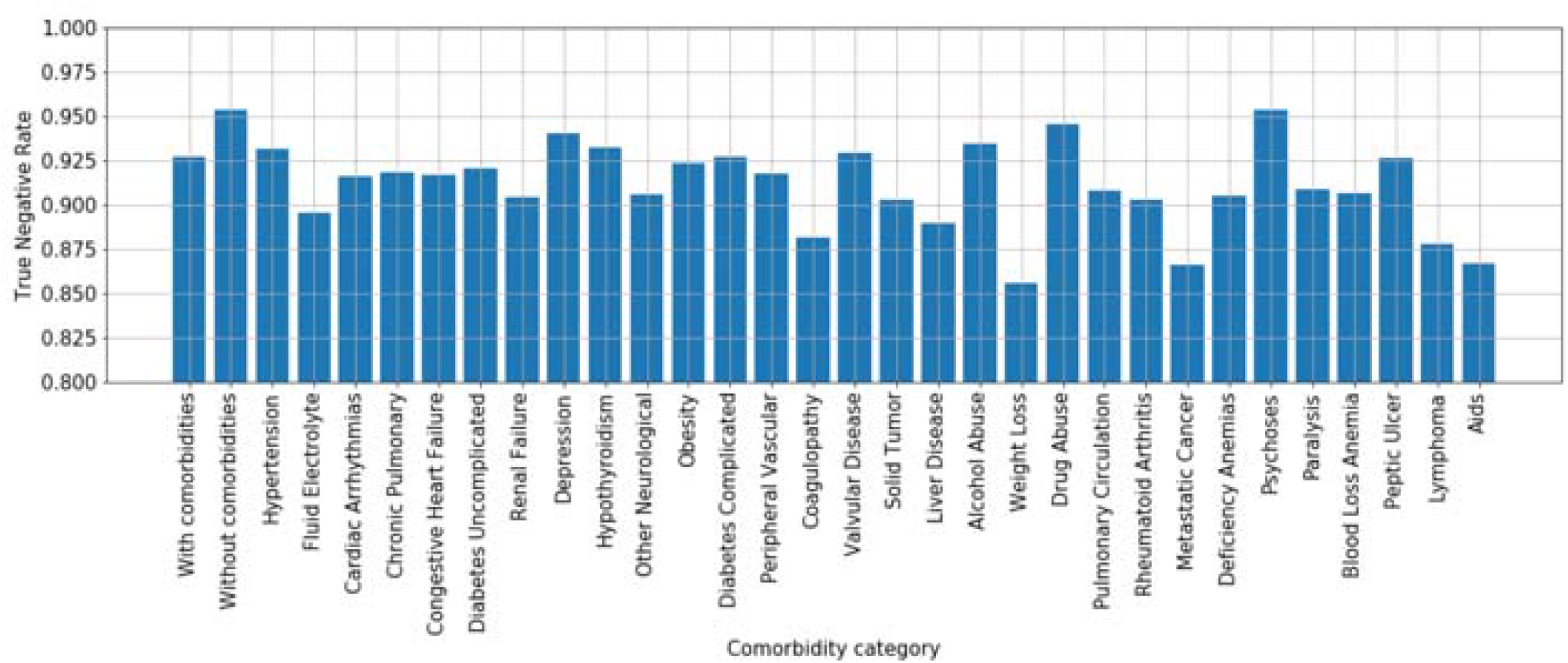
True negative rates (specificity) by comorbidity category. X-axis: “With comorbidities” - control patients with at least one comorbidity; “Without comorbidities” - control patients without any documented comorbidities; 30 comorbidity categories are ordered by prevalence shown in Figure 6 to highlight that the differences in True Negative Rate are not simple reflections of prevalence.

The model performs better at ruling out infection on control patients without comorbidities than those with comorbidities (TNR=0.95 vs. TNR=0.925), suggesting that confounding chronic conditions contribute to the false positive rate of the model. Interestingly, with respect to individual comorbidity categories, the model performs best at ruling out infection on control patients with neurological comorbidities (e.g., depression, psychoses), drug/alcohol abuse, and hypothyroidism; presumably since such conditions may have limited overlap in physiological biomarkers related to infection. The worst performing comorbidity categories include fluid/electrolyte disorders, coagulopathy, weight loss, metastatic cancer, lymphoma, anemia, and AIDS.

## DISCUSSION

Our work addresses the fundamental problem of early prediction of HAI, to allow prompt treatment and prevention of infectious disease transmission. We presented a large-scale, retrospective big data machine learning study that provides a data-driven approach to the problem, which can be tailored and adapted to different populations of interest. Infection can be detected by our model with high accuracy in its pre-symptomatic state at 48-hours before clinical suspicion.

Ensemble models proved to perform significantly better than both the established empirical rules and clinical scores, and logistic regression, with gradient boosting having the best performance. AdaBoost provided an interpretable model which allows us to map the feature importance to its relevance in clinical literature. For example, multiple laboratory values associated with inflammation ranked high in the feature importance metric, as well as features indicative of acidosis. High heart rate, high temperature and macro variability of vital signs were also indicative of infection, consistently with what has been reported in the literature [28] [29] [30] [31]. This characteristic of interpretability not only further validates our model, but also provides meaningful information in the clinical setting, quantifying the effect that appropriate action on each of these parameters would have in preventing HAI. It is well known that interpretability of the decision support model is vital to the acceptance of such a predictor in the clinical setting [33].

One important finding of our study is that the high performance of the model is obtained only by aggregating multiple biomarkers. No single “superfeature” exists that allows superior classification. This likely reflects at the same time the variable etiology of the HAI, which can be of different natures (respiratory, blood stream infection, sepsis, etc.), the individual variability in the response, and the multi-system nature of the effect of the infection on the patient’s physiology. On the other hand, it is still possible to obtain prediction performance that are clinically viable with a reasonable number of clinical measurements. We have showed that with a core set of 36 clinical measurements, the infection model performs at an AUC = 0.86 at one hour before clinical suspicion of infection.

The algorithm presented in this work could be implemented in a hospital setting by leveraging the existing monitoring systems and infrastructure. When risk of infection is predicted in advance, knowledge of the contributing parameters provided by the transparency of the model would allow secondary assessment and prompt intervention. While the best performing model employs a combination of laboratory test values and vital signs across 163 features, a model trained on 36 of the most frequently measured vital signs, labs and demographics achieves an AUC of 0.86 at 1-hour before clinical suspicion. Moreover, a model trained with only vital signs and demographics still achieves an acceptable area under the curve, equal to 0.81. A similar model could be employed in a context that is outside of the hospital (e.g. home monitoring via wearable devices) or in other situations where laboratory values are not easily obtainable.

## CONCLUSION

This study developed an algorithm for early identification of infection in hospitalized patients, using machine learning applied to large retrospective hospital datasets. The model is able to identify patients who are infected with reasonable performance up to 48 hours before clinical suspicion of infection (AUC > .85). The trained models utilize ensembles of decision trees, which are readily interpretable and provide ranked lists of feature importance. The primary model leveraging all available (163) vital signs, laboratory measurements and demographics achieves the best performance; however, a secondary model limited to the 36 most commonly measured clinical measurements still achieves an AUC=0.86 at 1-hour before clinical suspicion. The models compare favorably to established clinical rules and show high potential for real-world hospital deployment as a clinical decision support aid.

## Data Availability

MIMIC-III dataset is available in PhysioNet repository, https://mimic.physionet.org/. A portion of the eICU dataset used in this study is available in PhysioNet repository, https://eicu-crd.mit.edu; the remaining of the eICU dataset is proprietary to Philips.  The Banner Health dataset is a proprietary dataset that is not publicly shareable.

https://eicu-crd.mit.edu

## ACKNOWLEDGEMENTS

This work is sponsored by the US Department of Defense (DoD), Defense Threat Reduction Agency (DTRA) under project CB10560.

## LIST OF ABBREVIATIONS

HAI: Healthcare-associated infection
CDS: Clinical decision support
qSOFA: quick Sequential Organ Failure Assessment
SHAP: Shapley Additive exPlanations
Spec: Specificity
TNR: True Negative Rate
AUC: Area under the ROC curve

## DECLARATIONS

### Ethics approval and consent to participate

The MIMIC-III project was approved by the Institutional Review Boards of Beth Israel Deaconess Medical Center (Boston, MA) and the Massachusetts Institute of Technology (Cambridge, MA). Use of the eICU data was approved by the Philips Internal Committee for Biomedical Experiments. Banner Health data use was a part of an ongoing retrospective deterioration detection study approved by the Institutional Review Board of Banner Health and by the Philips Internal Committee for Biomedical Experiments.

Requirement for individual patient consent was waived because the project did not impact clinical care, was no greater than minimal risk, and all protected health information was removed from the limited dataset used in this study.

### Consent for publication

Not applicable

### Authors’ contributions

TF, CK, and BC participated in the study design, data preparation and analysis, machine learning model training, and contributed to writing of the manuscript. DN, SM, CZ, EG, and DM contributed to study design, data analysis, and contributed to writing of the manuscript. DS provided clinical consultation, manuscript review, and interpretation of results. JF provided clinical consultation and participated in hypothesis development, cohort identification, manuscript review and interpretation of results. All authors have read and approved the manuscript.

## ADDITIONAL FILES

Supplementary materials: Supplementary Material_v6.docx

^1^Oxygen Saturation is predominantly from pulse oximetry measurements and in addition blood gas measurements

