## Supplementary Material for "Machine Learning-based Clinical Decision Support for Infection Risk Prediction"

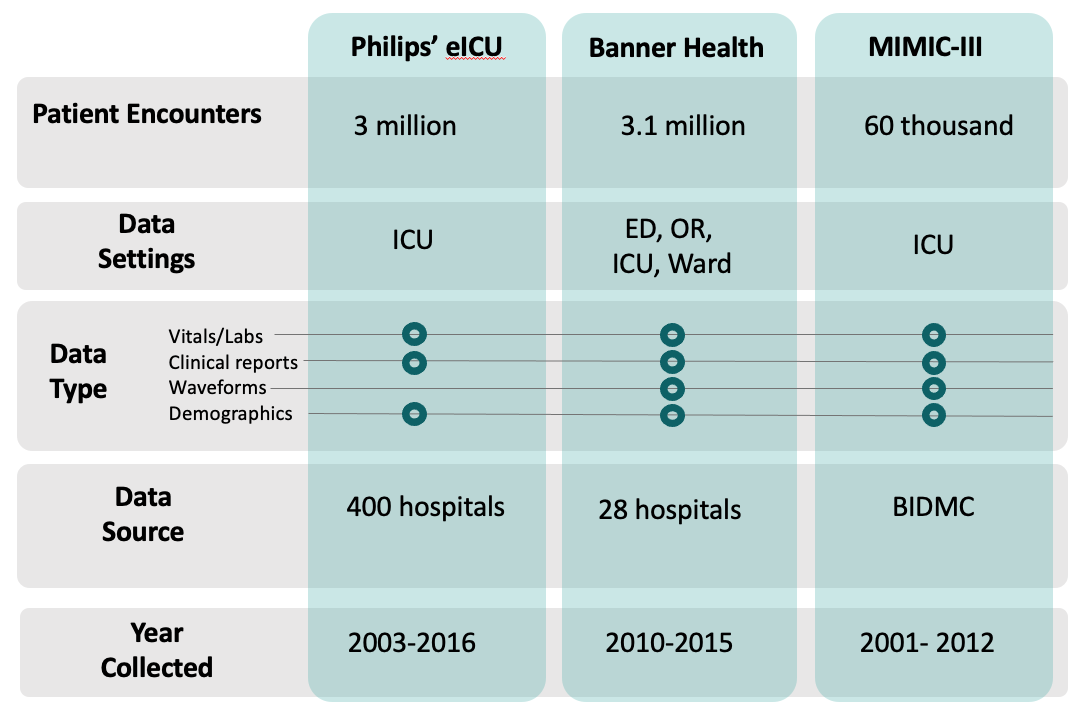

Figure S1: Data types present in each of the three datasets. ICU: Intensive Care Unit; ED: Emergency Department; OR: Operating Room; BIDMC: Beth Israel Deaconess Medical Center.

#### Generation of ICD-9 code set for infection diagnosis

Our patient encounters were before the full adoption of ICD-10, therefore we first curated a list of 479 ICD-9 (International Classification of Diseases, Ninth Revision) codes that are associated with the diagnoses of infection. These ICD-9 codes are extracted from 75 articles (see [1] and [2]) that were published between 1993 and 2015, where at least one type of healthcare-associated infection (HAI) was examined using a set of ICD-9 codes. For each type of HAI, we combined the ICD-9 codes used in the relevant publications. ICD-9 codes are present for the following HAI categories: bloodstream infection, sepsis, urinary system infection (USI), meningitis, aspergillosis, pneumonia, and bone, joint, tissue, soft tissue infection.

We further categorized the ICD-9 codes extracted from these articles into two classes: 1)

Infection Diagnosis – codes that directly identify the infection, and 2) Condition – codes that indicate associated symptoms or conditions of an infection. The Infection Cohort and the

Control Cohort are subsequently identified using the subset of 347 ICD-9 codes that are “Infection Diagnosis” codes.

#### Extraction of non-prophylactic antibiotic administration

We extracted non-prophylactic antibiotic administrations via examining administration route, durations and intervals. Specifically, we restricted antibiotic administration records to those that are delivered Intravenously (IV), and measured the intervals between each drug administration. Antibiotic administration records that are less than 24 hours apart are considered to be within the same treatment course. The duration of each treatment course is measured, and the ones that last more than 72 hours are considered as non-prophylactic treatment.

#### Comparison of cohort selection criteria

For a minority of patients from several eICU hospitals, the quality of reporting of microbiology tests did not allow the same cohort selection process, thus we employed an alternative technique using antibiotic administration records/ICD-9. This cohort selection criteria was validated using the MIMIC-III dataset by examining the similarity/discrepancy between cohorts identified via this method and via microbiology lab records/ICD-9 (Figure S2). We found that both infection and control cohorts defined by these criteria have large overlap with cohorts selected from the microbiology and ICD-9 criteria. Furthermore, antibiotic/ICD-9 criteria appear to be more conservative in selecting infection patients (11,734 infection patients) than microbiology/ICD-9 criteria (14,756), with 79% of identified infection patients also belonging to the infection cohort selected by microbiology lab/ICD-9. This result suggests that selecting infection and control patients using antibiotic/ICD-9 criteria is a valid approach.

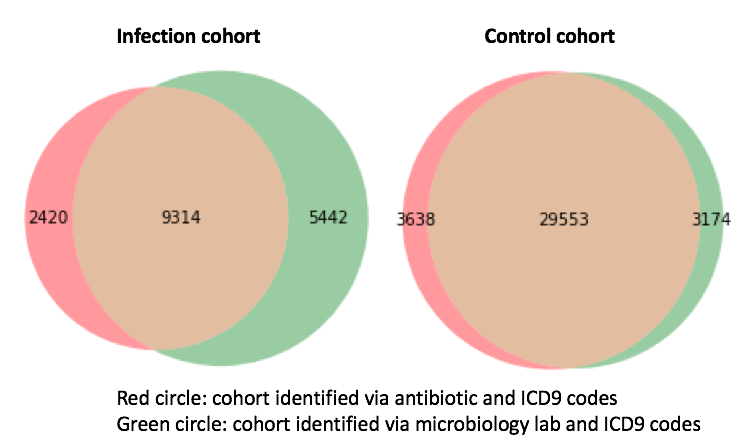

Figure S2: Overlap between cohort identified using antibiotics/ICD9 criteria (red circle) and using microbiology/ICD-9 criteria (green circle). Overlaid number shows the number of patients that is identified either exclusively by one criterion or by both.

#### Correcting for Dataset Heterogeneity and Covariate Shift

After features were extracted from each of the three hospital datasets, an extensive preprocessing and cleaning pipeline was applied to create a common and consistent dataset.

As a first pass, string matching routines were applied to identify and correct inconsistencies in concept naming conventions and feature measurement units between hospitals. In addition, the scripts parsed metadata to distinguish certain laboratory values by fluid sample (e.g., blood or urinary).

In a second pass, we applied machine learning techniques to identify additional sources of data heterogeneity and covariate shift. Specifically, each clinical measurement was used as an independent variable to train a classifier to predict which hospital dataset a given patient is from. We found that a few clinical measurements are highly predictive in classifying the hospital, indicating that the data distribution of these measurements are different across hospitals. We then manually inspected these features and identified the source of the problem, which typically fell into one of three categories:

1) Clinical measurements were missing in one of the datasets. The missing measurements were then re-extracted.
2) Clinical measurements were charted in different units, which were corrected manually.
3) Clinical measurements exhibited unknown covariate shift and/or could not be identified in one of the datasets. These included lipase and aPTT Ratio, which were excluded from further analyses.

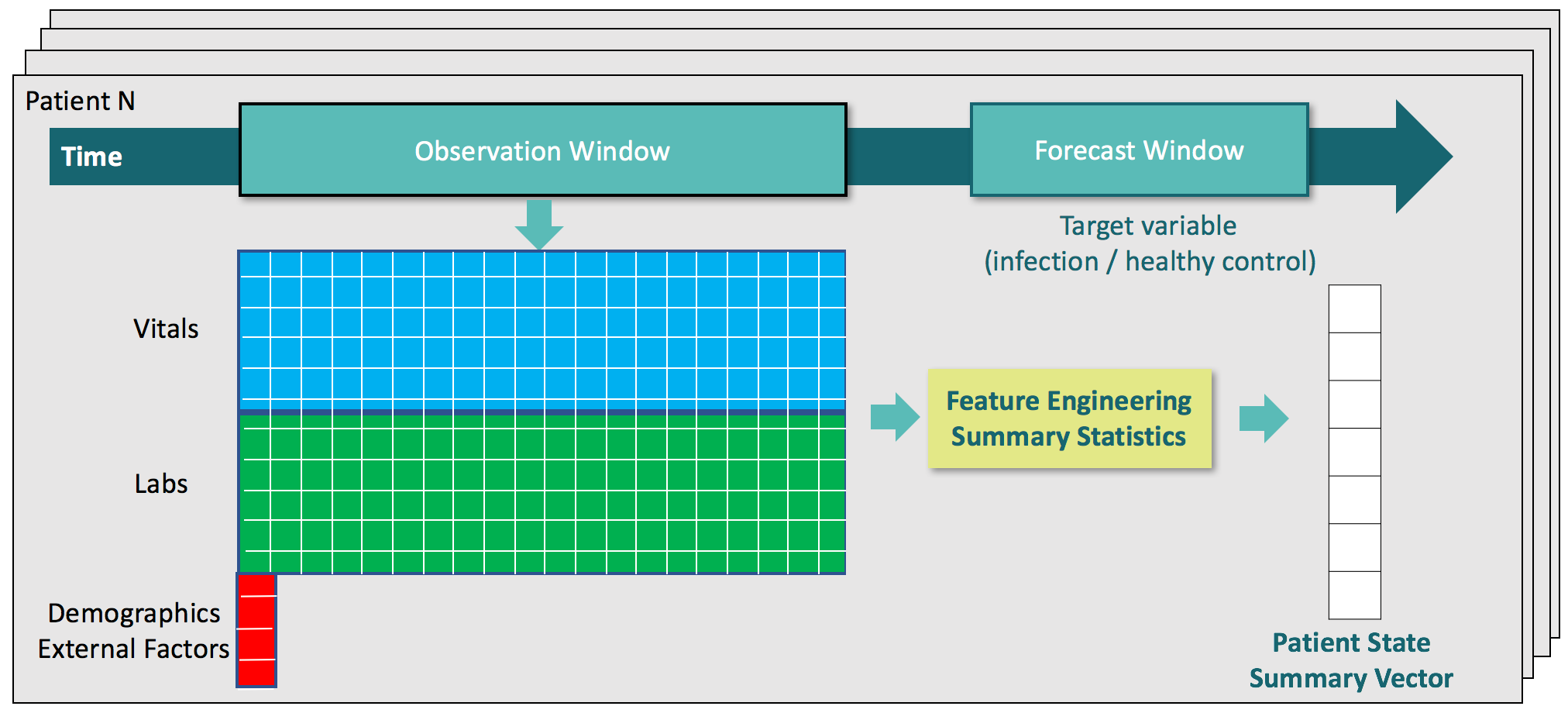

*Figure S3: Diagram of the feature extraction pipeline*

#### Imputation, Hyper parameter and performance evaluation

Analysis of healthcare data is typically challenged by incomplete and sparse data records. In our case, this resulted in missing values for features across patients. To rectify this, we employed Abstained Adaptive boosting algorithm [3] which abstains missing values from make predictions, Gradient Boosting of decision trees algorithm [4] which handles missing values by default, and used mean imputation for training logistic regression model.

Hyper parameter selection was performed using grid search. For logistic regression, Ridge Regularization parameters was chosen as the one who yield best average performance using cross-validation. For gradient boost, we finalized on depth of two trees as further increasing tree depth dose not increase performance substantially.

We evaluated performance using a number of metrics:

- Area under the ROC curve (AUC);
- Corrected Precision, or Corrected Positive Predictive Value (PPV(12.5%));
- True Positive Rate (Sensitivity, or Recall), including:
  - Sensitivity(Break-Even): Sensitivity at the break-even point, where Sensitivity and Precision are equal,
  - Sensitivity(80%): Sensitivity when Specificity=0.8,
  - Sensitivity(90%): Sensitivity when Specificity=0.9,
- True Negative Rate (Specificity).

We additionally applied the models that were trained on the one-hour before suspicion window to earlier time windows to characterize lead-time performance of the algorithm. When applying the model to earlier time windows, the infection prevalence tended to vary slightly because of lack of data for some infection patients at earlier time windows. Since precision (PPV) is sensitive to the prevalence, we report a corrected precision (PPV(12.5%)), which normalizes the PPV to the ambient infection prevalence (12.5%). This was done using Bayes’ rule that relates precision to sensitivity, specificity, and prevalence:

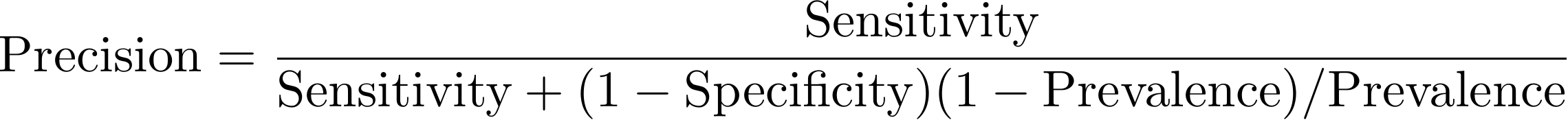

Specifically, a classifier’s corrected PPV(12.5%) was calculated by plugging its sensitivity and specificity metrics in the equation above and setting Prevalence=0.125.

Although the classifier decision threshold can be tuned to address different sensitivity/specificity tradeoffs, we report PPV(12.5%), Sensitivity, and Specificity metrics with respect to the break-even point, which identifies the decision threshold that roughly equalizes precision and sensitivity (recall).

### Bibliography

| [1] | O. T. J. A. Á. a. L. A. Redondo‐González, "Validity and Reliability of Administrative Coded Data for the Identification of Hospital‐Acquired Infections: An Updated Systematic Review with Meta‐Analysis and Meta‐Regression Analysis," *Health services research,* vol. 53, no. 3, pp. 1919-1956, 2018. |
| --- | --- |
| [2] | M. v. D. P. M. K. B. M. a. L. G. Van Mourik, "Accuracy of administrative data for surveillance of healthcare-associated infections: a systematic review," *BMJ open,* vol. 5, no. 8, p. p.e008424, 2015. |
| [3] | L. E. C. P. a. M. X.-W. B. Conroy, "A dynamic ensemble approach to robust classification in the presence of missing data," *Machine Learning,* pp. 443-463, 2016. |
| [4] | T. C. a. C. Guestrin, "Xgboost: A scalable tree boosting system," *22nd ACM SIGKDD international conference on knowledge discovery and data mining,* 2016. |

Table A‑1: Description of Extracted Features

| **Feature Name** | **Description** | **Units** |
| --- | --- | --- |
| Age | Age of the patient | Years |
| ALT (SGPT) | An alanine aminotransferase (ALT) test measures the amount of this enzyme in the blood. ALT is found mainly in the liver, but also in smaller amounts in the kidneys, heart, muscles, and pancreas. ALT was formerly called serum glutamic pyruvic transaminase (SGPT). ALT is measured to see if the liver is damaged or diseased. | Units/L |
| AST (SGOT) | An aspartate aminotransferase (AST) test measures the amount of this enzyme in the blood. AST is normally found in red blood cells, liver, heart, muscle tissue, pancreas, and kidneys. AST formerly was called serum glutamic oxaloacetic transaminase (SGOT). When body tissue or an organ such as the heart or liver is diseased or damaged, additional AST is released into the bloodstream. | Units/L |
| Albumin | An albumin test checks urine for a protein called albumin. Albumin is normally found in the blood and filtered by the kidneys. When the kidneys are working as they should, there may be a very small amount of albumin in the urine. But when the kidneys are damaged, abnormal amounts of albumin leak into the urine. This is called albuminuria. If the amount of albumin is very small, but still abnormal, it is called microalbuminuria. Albuminuria is most often caused by kidney damage from diabetes. But many other conditions can lead to kidney damage. These include high blood pressure, heart failure, cirrhosis, and lupus. | g/dL |
| Alkaline Phosphatase | An alkaline phosphatase (ALP) test measures the amount of the enzyme ALP in the blood. ALP is made mostly in the liver and in bone with some made in the intestines and kidneys. It also is made by the placenta of a pregnant woman. The liver makes more ALP than the other organs or the bones. Some conditions cause large amounts of ALP in the blood. These conditions include rapid bone growth (during puberty), bone disease (such as Paget's disease or cancer that has spread to the bones), a disease that affects how much calcium is in the blood (hyperparathyroidism), vitamin D deficiency, or damaged liver cells. | Units/L |
| Ammonia | An ammonia test measures the amount of ammonia in the blood. Most ammonia in the body forms when protein is broken down by bacteria in the intestines. The liver normally converts ammonia into urea, which is then eliminated in urine. Ammonia levels in the blood rise when the liver is not able to convert ammonia to urea. This may be caused by cirrhosis or severe hepatitis. | ug/dL |
| Amylase | An amylase test measures the amount of this enzyme in a sample of blood taken from a vein or in a sample of urine. Normally, only low levels of amylase are found in the blood or urine. But if the pancreas or salivary glands become damaged or blocked, more amylase is usually released into the blood and urine. In the blood, amylase levels rise for only a short time. In the urine, amylase may remain high for several days. | Units/L |
| Anion Gap | The anion gap (AG or AGAP) is a value calculated from the results of multiple individual medical lab tests. It may be reported with the results of an Electrolyte Panel, which is often performed as part of a Comprehensive Metabolic Panel. The anion gap is the difference between the measured cations (positively charged ions) and the measured anions (negatively charged ions) in serum, plasma, or urine. The magnitude of this difference (i.e., "gap") in the serum is often calculated in medicine when attempting to identify the cause of metabolic acidosis, a lower than normal pH in the blood. If the gap is greater than normal, then high anion gap metabolic acidosis is diagnosed. The term "anion gap" usually implies "serum anion gap", but the urine anion gap is also a clinically useful measure. | mEq/L |
| BNP | A brain natriuretic peptide (BNP) test measures the amount of the BNP hormone in your blood. BNP is made by your heart and shows how well your heart is working. Normally, only a low amount of BNP is found in your blood. But if your heart has to work harder than usual over a long period of time, such as from heart failure, the heart releases more BNP, increasing the blood level of BNP. | pg/mL |
| BUN | A blood urea nitrogen (BUN) test measures the amount of nitrogen in your blood that comes from the waste product urea. Urea is made when protein is broken down in your body. Urea is made in the liver and passed out of your body in the urine. A BUN test is done to see how well your kidneys are working. If your kidneys are not able to remove urea from the blood normally, your BUN level rises. Heart failure, dehydration, or a diet high in protein can also make your BUN level higher. Liver disease or damage can lower your BUN level. A low BUN level can occur normally in the second or third trimester of pregnancy. Blood urea nitrogen (BUN) and creatinine tests can be used together to find the BUN-to-creatinine ratio (BUN:creatinine). A BUN-to-creatinine ratio can help your doctor check for problems, such as dehydration, that may cause abnormal BUN and creatinine levels. | mg/dL |
| Bands | The count of less mature neutrophils -- a type of white blood cell (WBC; the major types of white blood cells are neutrophils, lymphocytes, monocytes, eosinophils, and basophils), from a blood count: the less mature neutrophils are known as "bands" or "stabs" because they are band or rod-like, instead of appearing segmented as the more mature neutrophils (called "segs"). Abnormal counts of neutrophils may indicate bacterial infection, and are seen in leukemias. They may also be raised in acute viral infections. | % (of WBC concentration) |
| Base Excess | The base excess is used for the assessment of the metabolic component of acid-base disorders, and indicates whether the person has metabolic acidosis or metabolic alkalosis. Contrasted with the bicarbonate levels, the base excess is a calculated value intended to completely isolate the non-respiratory portion of the pH change. | mEq/L |
| Basophils | The count of basophils - a type of white blood cell (WBC; the major types of white blood cells are neutrophils, lymphocytes, monocytes, eosinophils, and basophils), from a blood count. Each type of cell plays a different role in protecting the body. The numbers of each one of these types of white blood cells give important information about the immune system. Too many or too few of the different types of white blood cells can help find an infection, an allergic or toxic reaction to medicines or chemicals, and many conditions, such as leukemia. | % (of WBC concentration) |
| Bicarbonate | In diagnostic medicine, the blood value of bicarbonate is one of several indicators of the state of acid–base physiology in the body. It is measured, along with carbon dioxide, chloride, potassium, and sodium, to assess electrolyte levels in an electrolyte panel test. | mEq/L |
| CPK | A creatine kinase (CK) test (also called creatine phosphokinase (CPK), or phosphocreatine kinase) checks the level of the enzyme creatine kinase, which is found in heart tissue and skeletal muscles. This enzyme also can be found in smaller amounts in the brain. A blood test to check the level of CK can show if there has been damage to the heart, skeletal muscles, brain, and sometimes other parts of the body. CK is made up of three smaller types of enzymes, called isoenzymes: MM, MB, and BB. A doctor looks not only at the total level of CK but also at the level of these smaller parts to find a health problem. CK might be used to help diagnose a heart attack. | Units/L |
| CPK-MB | The CPK-MB test (one of three subtypes of CPK) is a cardiac marker used to assist diagnoses of an acute myocardial infarction. It measures the blood level of CK-MB (creatine kinase-muscle/brain), the bound combination of two variants (isoenzymes CKM and CKB) of the enzyme phosphocreatine kinase. In some locations, the test has been superseded by the troponin test. However, recently, there have been improvements to the test that involve measuring the ratio of the CK-MB1 and CK-MB2 isoforms. The newer test detects different isoforms of the B subunit specific to the myocardium whereas the older test detected the presence of cardiac-related isoenzyme dimers. Many cases of CK-MB levels exceeding the blood level of total CK have been reported, especially in newborns with cardiac malformations, especially ventricular septal defects. This reversal of ratios is in favor of pulmonary emboli or vasculitis. An autoimmune reaction creating a complex molecule of CK and IgG should be taken into consideration. | ng/mL |
| CPK-MB Index | Ratio of CK-MB to total CK (see CPK-MB for details) | % |
| CRP | A C-reactive protein (CRP) test is a blood test that measures the amount of a protein called C-reactive protein in your blood. C-reactive protein measures general levels of inflammation in your body. High levels of CRP are caused by infections and many long-term diseases. But a CRP test cannot show where the inflammation is located or what is causing it. Other tests are needed to find the cause and location of the inflammation. | mg/dL |
| CVP | The central venous pressure (CVP) is the pressure measured in the central veins close to the heart. It indicates mean right atrial pressure and is frequently used as an estimate of right ventricular preload. The CVP does not measure blood volume directly, although it is often used to estimate this. | mmHg |
| Calcium | A test for calcium in the blood checks the calcium level in the body that is not stored in the bones. Calcium is the most common mineral in the body and one of the most important. The body needs it to build and fix bones and teeth, help nerves work, make muscles squeeze together, help blood clot, and help the heart to work. Almost all of the calcium in the body is stored in bone. Normally the level of calcium in the blood is carefully controlled. When blood calcium levels get low (hypocalcemia), the bones release calcium to bring it back to a good blood level. When blood calcium levels get high (hypercalcemia), the extra calcium is stored in the bones or passed out of the body in urine and stool. The amount of calcium in the body depends on the amount of: Calcium you get in your food; Calcium and vitamin D your intestines absorb. Phosphate in the body; and Certain hormones, including parathyroid hormone, calcitonin, and estrogen in the body. | mg/dL |
| Carboxyhemoglobin | Carboxyhemoglobin or carboxyhaemoglobin (symbol COHb or HbCO) is a stable complex of carbon monoxide and hemoglobin (Hb) that forms in red blood cells upon contact with carbon monoxide (CO). Exposure to small concentrations of CO hinder the ability of Hb to deliver oxygen to the body, because carboxyhemoglobin forms more readily than does oxyhemoglobin (HbO2). CO is produced in normal metabolism and is also a common chemical. Tobacco smoking (through carbon monoxide inhalation) raises the blood levels of COHb by a factor of several times from its normal concentrations. | % |
| Cardiac Output | Cardiac output is a term used in cardiac physiology that describes the volume of blood being pumped by the heart, in particular by the left or right ventricle, per unit time. Cardiac output is the product of the heart rate (HR), or the number of heart beats per minute (bpm), and the stroke volume (SV), which is the volume of blood pumped from the ventricle per beat; thus, CO = HR × SV. Values for cardiac output are usually denoted as L/min. For a healthy person weighing 70 kg, the cardiac output at rest averages about 5 L/min; assuming a heart rate of 70 beats/min, the stroke volume would be approximately 70 mL. Because cardiac output is related to the quantity of blood delivered to various parts of the body, it is an important indicator of how efficiently the heart can meet the body's demands for perfusion. | L/min |
| Chloride | A chloride test measures the level of chloride in your blood or urine. Chloride is one of the most important electrolytes in the blood. It helps keep the amount of fluid inside and outside of your cells in balance. It also helps maintain proper blood volume, blood pressure, and pH of your body fluids. Tests for sodium, potassium, and bicarbonate are usually done at the same time as a blood test for chloride. Most of the chloride in your body comes from the salt (sodium chloride) you eat. Chloride is absorbed by your intestines when you digest food. Extra chloride leaves your body in your urine. Sometimes a test for chloride can be done on a sample of all your urine collected over a 24-hour period (called a 24-hour urine sample) to find out how much chloride is leaving your body in your urine. Chloride can also be measured in skin sweat to test for cystic fibrosis. | mEq/L |
| Cortisol | A cortisol test is done to measure the level of the hormone cortisol in the blood. The cortisol level may show problems with the adrenal glands or pituitary gland. Cortisol is made by the adrenal glands. Cortisol levels go up when the pituitary gland releases another hormone called adrenocorticotropic hormone (ACTH). Cortisol has many functions. It helps the body use sugar (glucose) and fat for energy (metabolism), and it helps the body manage stress. Cortisol levels can be affected by many conditions, such as physical or emotional stress, strenuous activity, infection, or injury. Normally, cortisol levels rise during the early morning hours and are highest about 7 a.m. They drop very low in the evening and during the early phase of sleep. But if you sleep during the day and are up at night, this pattern may be reversed. If you do not have this daily change (diurnal rhythm) in cortisol levels, you may have overactive adrenal glands. This condition is called Cushing's syndrome. The timing of the cortisol test is very important because of the way cortisol levels vary throughout a day. If your doctor thinks you might make too much cortisol, the test will probably be done late in the day. If your doctor thinks you may not be making enough, a test is usually done in the morning. | ug/dL |
| Creatinine | Creatinine and creatinine clearance tests measure the level of the waste product creatinine in your blood and urine. These tests tell how well your kidneys are working. Another substance, creatine, is formed when food is changed into energy through a process called metabolism. Creatine is broken down into creatinine. Your kidneys take creatinine out of your blood and pass it out of your body in urine. If your kidneys are damaged and can't work as they should, the amount of creatinine in your urine goes down while its level in your blood goes up. Three types of tests can be done: Blood creatinine level; Creatinine clearance (how well creatinine is removed from your blood by your kidneys; the test is done on both a blood sample and on a sample of urine collected over 24 hours; and Blood urea nitrogen-to-creatinine ratio (BUN:creatinine). | mg/dL |
| Digoxin | A digoxin test is a blood test that your doctor can use to determine the level of the medication digoxin in your blood. Digoxin is a drug that contains cardiac glycosides. People take it to treat heart failure and irregular heartbeats. Digoxin is available in oral form. Your body absorbs it, and it then travels to your body’s tissues, especially your heart, kidney, and liver. Your doctor performs digoxin testing to make sure that you aren’t receiving too much or too little of the drug. Your doctor should monitor the level of digoxin in your blood because the drug has a narrow safe range. Digoxin, sold under the brand name Lanoxin among others, is a medication used to treat various heart conditions. Most frequently it is used for atrial fibrillation, atrial flutter, and heart failure. Digoxin is taken by mouth or by injection into a vein. | ng/mL |
| Direct Bilirubin | A bilirubin test measures the amount of bilirubin in a blood sample. Bilirubin is a brownish yellow substance found in bile. It is produced when the liver breaks down old red blood cells. Bilirubin is then removed from the body through the stool (feces) and gives stool its normal color. Bilirubin circulates in the bloodstream in two forms: (1) Indirect (or unconjugated) bilirubin. This form of bilirubin does not dissolve in water (it is insoluble). Indirect bilirubin travels through the bloodstream to the liver, where it is changed into a soluble form (direct or conjugated); and (2) Direct (or conjugated) bilirubin. Direct bilirubin dissolves in water (it is soluble) and is made by the liver from indirect bilirubin. Total bilirubin and direct bilirubin levels are measured directly in the blood, whereas indirect bilirubin levels are derived from the total and direct bilirubin measurements. When bilirubin levels are high, the skin and whites of the eyes may appear yellow (jaundice). Jaundice may be caused by liver disease (hepatitis), blood disorders (hemolytic anemia), or blockage of the tubes (bile ducts) that allow bile to pass from the liver to the small intestine. Mild jaundice in newborns usually does not cause problems. But too much bilirubin (hyperbilirubinemia) in a newborn baby can cause brain damage (kernicterus) and other serious problems. So some babies who develop jaundice may need treatment to lower their bilirubin levels. | mg/dL |
| ESR | An erythrocyte sedimentation rate (ESR or sed rate) test helps identify diseases that cause inflammation, such as polymyalgia rheumatica. ESR is the rate at which red blood cells sediment in a period of one hour. It is a common hematology test, and is a non-specific measure of inflammation. To perform the test, anticoagulated blood is traditionally placed in an upright tube, known as a Westergren tube, and the rate at which the red blood cells fall is measured and reported in mm at the end of one hour. | mm/hr |
| End Tidal CO2 | End-tidal carbon dioxide (ETco2) monitoring provides valuable information about CO2 production and clearance (ventilation). Also ETco2 is called as capnometry or capnography. This noninvasive technique provides a breath-by-breath analysis and a continuous recording of ventilatory status. In fact, it’s commonly called the “ventilation vital sign.” By providing instantaneous feedback on the patient’s ventilation effectiveness, ETco2 monitoring gives early warning of respiratory compromise. It also may reflect cardiac perfusion changes and has been used to indicate the effectiveness of chest compressions in cardiac arrest. What’s more, it confirms endotracheal tube placement and helps monitor ventilator circuit integrity. A standard of care in the operating room for more than 25 years, ETco2 monitoring is becoming a common adjunct in the intensive-care and procedural-care settings. In the prehospital arena, it provides immediate feedback on the patient’s ventilatory status. | mmHg |
| Eosinophils | The count of basophils -- a type of white blood cell (WBC; the major types of white blood cells are neutrophils, lymphocytes, monocytes, eosinophils, and basophils), from a blood count. Each type of cell plays a different role in protecting the body. The numbers of each one of these types of white blood cells give important information about the immune system. Too many or too few of the different types of white blood cells can help find an infection, an allergic or toxic reaction to medicines or chemicals, and many conditions, such as leukemia. | % (of WBC concentration) |
| Fe (Iron) | An iron test checks the amount of iron in the blood to see how well iron is metabolized in the body. Iron (Fe) is a mineral needed for hemoglobin, the protein in red blood cells that carries oxygen. Iron is also needed for energy, good muscle and organ function. About 70% of the body's iron is bound to hemoglobin in red blood cells. The rest is bound to other proteins (transferrin in blood or ferritin in bone marrow) or stored in other body tissues. When red blood cells die, their iron is released and carried by transferrin to the bone marrow and to other organs such as the liver and spleen. In the bone marrow, iron is stored and used as needed to make new red blood cells. | ug/dL |
| Ferritin | A ferritin blood test checks the amount of ferritin in the blood. Ferritin is a protein in the body that binds to iron; most of the iron stored in the body is bound to ferritin. Ferritin is found in the liver, spleen, skeletal muscles, and bone marrow. Only a small amount of ferritin is found in the blood. The amount of ferritin in the blood shows how much iron is stored in your body. | ng/mL |
| FiO2 | Fraction of inspired oxygen | % |
| Fibrinogen | Fibrinogen is a blood plasma protein that is a thrombin precursor and therefore an indicator of blood coagulation and clotting. | mg/dL |
| Folate | Vitamin B9, broadly required for normal hematology and other physiological processes. | ng/mL |
| Free T4 | T4 is Thyroxine, a thyroid hormone used clinically to check thyroid function. Free indicates not bound. | ng/dL |
| Gender | Gender of the patient | N/A |
| Glucose | Simple sugar required for metabolism, a clinical measure used to assess metabolism and diabetes | mg/dL |
| HDL | High density lipoprotein, one of several clinical indicators of cholesterol metabolism | mg/dL |
| Haptoglobin | Protein that removes free hemoglobin from plasma, used clinically as a marker of hemolysis, i.e., lysis of RBCs | mg/dL |
| Heart Rate | Clinical measure of heart beat frequency | Bpm |
| Height | Height of the patient | cm |
| Hematocrit | Volume fraction of red blood cells to whole blood (including cells and plasma) | % |
| Hemoglobin | Protein that transports O2 in the blood | g/dL |
| ICP | Intercranial pressure, the internal pressure inside the skull | mmHg |
| Invasive BP Diastolic | Blood pressure when heart is relaxed as measured by inserting a cannula into an artery | mmHg |
| Invasive BP Mean | Mean of the invasive systolic and diastolic BP | mmHg |
| Invasive BP Systolic | Blood pressure when heart is contracted as measured by inserting a cannula into an artery | mmHg |
| Ionized Calcium | Free calcium in the blood, essential for many organ systems | mmol/L |
| LDH | Lactic Acid Dehydrogenase, an enzyme involved in metabolism | Units/L |
| LDL | Low density lipoprotein, one of several clinical indicators of cholesterol metabolism. | mg/dL |
| LPM O2 | Flowrate of oxygen a patient receives in liters per minute (LPM) | L/min |
| Lactate | Metabolic byproduct with multiple clinical implications | mmol/L |
| Lipase | Pancreatic enzyme that catalyzes breakdown of fatty acids, used to monitor pancreatic health | Units/L |
| Lymphocytes | A measure of the amount of lymphocytes, part of the whole blood cell (WBS) differential test | % (of WBC concentration) |
| MCH | Mean cell hemoglobin, the calculated amount of hemoglobin per red blood cell | pg |
| MCHC | Mean corpuscular hemoglobin concentration, the calculated amount of hemoglobin per average volume of red blood cell. | g/dL |
| MCV | Mean corpuscular volume. Average red blood cell volume | fL |
| MPV | Mean platelet volume | fL |
| Magnesium | Amount of magnesium in the blood | mg/dL |
| Methemoglobin | Methemoglobin is an abnormal form of hemoglobin that has a different O2 affinity. Large amount can have multiple implications. The measurement of methemoglobin is not a part of standard CBC. | % |
| Monocytes | A type of white blood cell, part of innate immunity | % (of WBC concentration) |
| Myoglobin | Myoglobin stores O2 in muscle tissue and its amount can indicate muscle disorders or damage. | ng/mL |
| Neutrophils | A type of white blood cell, part of innate immunity | % (of WBC concentration) |
| Noninvasive BP Diastolic | Blood pressure in relaxation phase of cardiac cycle, measured non-invasively | mmHg |
| Noninvasive BP Mean | Blood pressure averaged over cardiac cycle, measured non- invasively | mmHg |
| Noninvasive BP Systolic | Blood pressure in contraction phase of cardiac cycle, measured non-invasively | mmHg |
| Oxyhemoglobin | Saturation of oxygen in hemoglobin | % |
| PAOP | Pulmonary artery diastolic pressure | mmHg |
| PA Diastolic | Pulmonary artery mean pressure | mmHg |
| PA Mean | Pulmonary artery systolic pressure | mmHg |
| PA Systolic | Pulmonary artery wedge pressure | mmHg |
| PEEP | Positive end expiratory pressure – a setting on ventilator | cmH20 |
| PT | Prothrombin time measuring the time to clot | sec |
| PT-INR | Prothrombin time measuring the time to clot- International Normalized Ratio | ratio |
| PTT | Partial thromboplastin time - time taken for blood to clot | sec |
| aPTT Ratio | Derivative of the activated partial thromboplastin time | ratio |
| PVR | Pulmonary vascular resistance - (https://www.ncbi.nlm.nih.gov/pmc/articles/PMC6132188/) a calculated value. Requires a pulmonary artery catheter |  |
| PVRI | Pulmonary vascular resistance index -(https://www.ncbi.nlm.nih.gov/pmc/articles/PMC6132188/) a calculated value. Requires a pulmonary artery catheter |  |
| PaCO2 | Partial pressure carbon dioxide - part of ABG test | mmHg |
| PaO2 | Partial pressure oxygen - part of ABG test | mmHg |
| Phenytoin | Anti-convulsant medication | ug/mL |
| Phosphate | Level of phosphate in blood | mg/dL |
| Phosphorous | Level of phosphorous in blood | mg/dL |
| Platelets | Number of platelets in blood | K/uL |
| Potassium | Level of potassium in blood | mEq/L |
| Prealbumin | Prealbumin level in blood | mg/dL |
| Pressure Support | Pressure support ventilation (setting) | cmH20 |
| RBC | Red blood cell count | M/uL |
| RDW | Red blood cell distribution width | % |
| Respiration | Spontaneous respiration rate | bpm |
| ST1 | Elevation of ST segment - https://github.com/MIT-LCP/eicu-website/issues/39 |  |
| ST2 | Elevation of ST segment - https://github.com/MIT-LCP/eicu-website/issues/39 |  |
| ST3 | Elevation of ST segment - https://github.com/MIT-LCP/eicu-website/issues/39 |  |
| SVR | Hemodynamic parameters - Systemic Vascular Resistance SVR = 80*(MAP-RAP)/CO where Cardiac Output CO= HR*SV/1000 |  |
| SVRI | Hemodynamic parameters - Systemic Vascular Resistance Index SVRI = 80 * (MAP-RAP)/CI where Cardiac Index CI = CO/BSA |  |
| SpO2 | This measures the percentage of how much hemoglobin is saturated with oxygen. | % |
| Sodium | Blood sodium level from basic metabolic panel | mEq/L |
| T4 | Free thyroxine (free T4) tests are used to help evaluate thyroid function and diagnose thyroid diseases, including hyperthyroidism and hypothyroidism, usually after discovering that the thyroid stimulating hormone level is abnormal. | ug/dL |
| TIBC | Total iron binding capacity (TIBC) is a blood test to see if you have too much or too little iron in your blood. | ug/dL |
| TSH | The level of thyroid that stimulates hormone in blood | mcU/mL |
| Temperature | Temperature of the patient | C |
| Total Bilirubin | Bilirubin in the blood - a measurement of liver function | mg/dL |
| Total CO2 | Total amount of CO2 - sum of HCO3 and PCO2 in blood gas test | mEq/L |
| Total Cholesterol | Total cholesterol in blood | mg/dL |
| Total Protein | Total protein in the blood - total amount of albumin and globulin; part of complete metabolic panel test | g/dL |
| Transferrin | Transferrin is the main protein in the blood that binds to iron and transports it throughout the body. | mg/dL |
| Triglycerides | Triglycerides are a type of fat (lipid) found in blood. The body converts any calories it doesn't need to use right away into triglycerides. | mg/dL |
| Troponin – I | The level of Troponin I protein in the blood, typically released when heart muscle has been damaged | ng/mL |
| Troponin – T | The level of Troponin T protein in the blood, typically released when heart muscle has been damaged | ng/mL |
| Uric Acid | Serum uric acid measurement, which determines how much uric acid is present in the blood | mg/dL |
| Urinary Calcium | Calcium level in urine | mg/dL |
| Urinary Chloride | Chloride level in urine | mEq/L |
| Urinary Creatinine | Creatinine level in urine | mg/dL |
| Urinary Potassium | Potassium level in urine | mmol/L |
| Urinary RBC | Red blood cell count in urine | #/hpf |
| Urinary Sodium | Sodium level in urine | mEq/L |
| Urinary Specific Gravity | A measurement of urine concentration, which compares the amount of substances dissolved in urine as compared to pure water |  |
| Urinary WBC | White blood cell count in urine | #/hpf |
| Urinary pH | pH in urine, part of urinalysis |  |
| Vancomycin Trough | The lowest amount of vancomycin in blood, typically drawn 1-2 hours before the next dose | ug/mL |
| Vitamin B12 | Vitamin B12 level in blood - B12 is important for brain health, blood cell production, and proper nerve function. | pg/mL |
| WBC | White blood cell count | K/uL |
| Weight | Weight of the patient | kg |
| pH | pH in arterial blood gas test |  |
